# Predictors of eviction and quality of tenant-landlord relationships during the 2020-2021 eviction moratorium in the U.S.

**DOI:** 10.1101/2021.08.27.21262736

**Authors:** Jack Tsai, Minda Huang, John R. Blosnich, Eric B. Elbogen

## Abstract

In 2020, the Centers for Disease Control and Prevention (CDC) issued several agency orders that put into effect a national moratorium on evictions for over one year to limit transmission of Coronavirus Disease 2019 (COVID-19). Little is known about landlord and tenant behaviors during the eviction moratorium. The current study used three waves of data from May 2020-April 2021 from a nationally representative sample of U.S. middle- and low-income tenants (n= 3,393 in Wave 1, n= 1,311 in Wave 2, and 814 in Wave 3) to examine tenants who were evicted during the eviction moratorium and the reported effects of the moratorium on tenant rental payments and tenant-landlord relationships. Across three Waves, 4.3% of tenants reported experiencing an eviction during the moratorium and 6-23% of tenants reported delaying paying rent because of the moratorium. Multivariable analyses found that tenants who delayed paying their rent, were female, or had a history of mental illness or substance use disorder were significantly more likely to report the eviction moratorium had a negative effect on the relationship with their landlord. Analyses also revealed that testing positive for COVID-19 was not a significant predictor of eviction but tenants with a history a homelessness were more than 9 times as likely to report an eviction than those without such a history. Together, these findings suggest the eviction moratorium has had some unintended consequences on rent payments and tenant-landlord relationships that need to be considered in the aftermath of the COVID-19 pandemic.

---

The Coronavirus Disease 2019 (COVID-19) pandemic was an extraordinary event that resulted in mass social distancing and temporary closing of businesses across the U.S. to minimize virus transmission. In late March 2020, Congress passed the Coronavirus Aid, Relief, and Economic Security (CARES) Act (P.L. 116-136) including Section 4024 which provided a temporary moratorium on evictions nationally for the next 4 months (McCarty & Carpenter, 2020). Then in September 4, 2020, the Centers for Disease Control and Prevention (CDC) issued the agency order entitled “Temporary Halt in Residential Evictions To Prevent Spread of COVID-19” that put into effect a federal eviction moratorium thru December 31, 2020 during which time “a landlord, owner of a residential property, or another person with a legal right to pursue eviction or possessory action, shall not evict any covered persons from any residential property in any jurisdiction to which the order applies” (Centers for Disease Control and Prevention, 2020). The order was then extended twice lasting until June 30, 2021 (Centers for Disease Control and Prevention, 2021c). Upon expiration of the order, the CDC issued a new eviction moratorium on August 3, 2021 for “communities with substantial or high levels of community transmission of COVID-19” (Centers for Disease Control and Prevention, 2021d).

Following the CDC’s initial order, studies began to emerge that supported the order with findings that evictions could increase COVID-19 virus transmission. One study found that across the U.S., local policies that limited evictions reduced COVID-19 infections by 3.8% and deaths by 11% (Jowers et al., 2021). Another study modeled the effect of evictions on COVID-19 transmission under two counterfactual scenarios: one in which an eviction moratorium was enforced and the other in which evictions were allowed to resume (Nande et al., 2021). The study found that across scenarios, evictions led to significant increases in COVID-19 infections and this increase was especially profound among low-income populations. While these studies provided evidence of the effect of evictions on COVID-19, there has been little study of the reverse—the effect of COVID-19 on evictions—which is the focus of this study.

Many individuals infected with COVID-19 experienced only temporary flu-like symptoms, but some individuals experienced illness that was severe enough to require hospitalization or resulted in death (Khalili et al., 2020). Regardless of symptom severity, CDC issued guidelines that individuals who tested positive for COVID-19 were to isolate themselves for 10-14 days (Centers for Disease Control and Prevention, 2021b). Isolation procedures may have disrupted the lives of people with COVID-19 in terms of their employment, financial situation, and interpersonal relationships which may have put them at greater risk for eviction. Some individuals with severe symptoms of COVID-19 were debilitated, hospitalized, and took extended periods of time for recovery which further disrupted their lives and put them at risk for eviction. Extenuating circumstances may have been exacerbated by the psychological distress and substance abuse commonly reported during the pandemic (Tsai et al., 2021; Vindegaard & Benros, 2020). However, there are gaps in knowledge of landlord and tenant behaviors during this time, such as how often landlords still filed evictions despite the moratorium, how often tenants withheld rent because of the moratorium, and the effect of the moratorium on tenant-landlord relationships.

To address some of these gaps, we used a nationally representative sample of middle- and low-income U.S. tenants to: 1) examine characteristics of tenants who were evicted during the eviction moratorium; 2) analyze reported effects of the moratorium on rental payments and relationships with landlords; and 3) identify tenant characteristics associated with negative tenant-landlord relationships during the moratorium.

## Methods

Data were from three waves of surveys collected from a national sample of middle- and low-income U.S. adults to track health and social well-being during the COVID-19 pandemic. In Wave 1 (May-June 2020), 6,607 adults were enrolled and participated in the study. In Wave 2, these participants were followed up after 3-months (Sept-October 2020) with 3,169 participants retained. In Wave 3, these participants were again followed up after 6 months (February-April 2021) and 2,124 participants were retained. All three Waves occurred during the national eviction moratorium.

Eligibility criteria for the study were adults who were at least 22 years old, living in the U.S., and reported an annual personal gross income of $75,000 or less. A total of 9,760 individuals initially agreed to participate, but 6,607 (67.7%) met eligibility criteria in Wave 1, fulfilled validity checks, and completed the baseline assessment. This study focused on participants who were renting apartments (i.e., were tenants) during the study period including 3,393 tenants in Wave 1 (51.4% of total Wave 1 sample); 1,311 tenants in Wave 2 (41.4% of total Wave 2 sample); and 814 tenants in Wave 3 (38.3% of total Wave 3 sample).

Participants were recruited and compensated through Amazon Mechanical Turk (MTurk), an online labor market with over 500,000 participants across 200 countries that has become a popular method for conducting surveys and online interventions. To ensure data quality, only participants who had completed ≥50 approved previous Human Intelligence Tasks (HITs) and had an HIT approval rating ≥50% were invited. Cross-sample investigations have demonstrated that data obtained from MTurk is the same level of quality or higher than data collected from traditional subject pools such as community samples, college students, and professional panels especially when eligibility requirements and validity checks are implemented (Kees et al., 2017).

To maximize generalizability of our findings, we used raking procedures to create sample weights representative of the U.S. middle- and low-income adult population using data from 2018 American Community Survey to compute poststratification weights with respect to age, sex, race, ethnicity, and geographic region. All study procedures were approved by the institutional review board at the University of Texas Health Science Center at Houston.

### Measures

Sociodemographic information was collected through a self-report questionnaire. History of homelessness was defined as “did not have a stable night-time residence (such as living in streets, shelters, cars, etc.). COVID19 status was assessed by asking participants whether they have been tested for COVID19 and what the outcome was (i.e., positive, negative, not tested). They were also asked whether anyone close to them (e.g., friends, family) had tested positive for COVID19.

A number of eviction-related variables was assessed. Across all three Waves, participants were asked: “Are you currently at risk of being evicted?” and “Are you aware there has been a moratorium/ban on evictions?” with Yes/No response options for both questions. Participants were further asked, “How has the moratorium on evictions affected your relationship with your landlord” with response options ranging from -2 (Worsened the Relationship) to 2 (Improved the Relationship Greatly) and a “N/A or I don’t know” option as well. Participants were also asked: “Have you delayed paying your rent because of the moratorium/ban on eviction” with Yes/No and N/A response options. In Waves 2 and 3, participants were asked: “In the past 3 months, have you been evicted from an apartment?” with yes/no response options.

Social connectedness was assessed with the Medical Outcomes Study (MOS) Social Support Survey-Short Form(Holden et al., 2014) and a question about the number of close friends and relatives that participants had.

Physical health status was assessed by asking participants whether they’ve ever been diagnosed with any of 22 different physical health conditions (e.g., cancer, heart disease, arthritis) and the total number was summed.(Thomas et al., 2017)

Psychiatric history was assessed by asking participants whether they have ever been diagnosed with any of nine mental health or substance use disorders. Current mental health and substance use was assessed with validated measures, including the Patient Health Questionnaire (PHQ-4);(Kroenke et al., 2001) assessment of any past 2-week suicidal ideation using an item from the Mini-International Neuropsychiatric Interview (MINI)(Sheehan et al., 1998); assessment of any COVID-19 era-related stress using the Posttraumatic Stress Disorder Checklist for the Diagnostic and Statistical Manual for Mental Disorders, Fifth Edition (PCL-5)(Weathers et al., 2013) with the COVID-19 pandemic as an index stressor event; and assessment of substance abuse using the Alcohol Use Disorders Identification Test (AUDIT-C)(Bush et al., 1998) and questions about current use of cigarettes, vaping, and any illicit drugs in the past month.

### Data Analysis

First, descriptive statistics were calculated to examine the proportion of tenants who were at-risk of eviction, experienced an eviction, was aware of the federal eviction moratorium, and delayed paying rent because of the moratorium at each Wave. Tenants’ mean rated effect of the moratorium on their relationship with their landlord was also calculated for each Wave. The full sample of tenants at each Wave was used to maximize use of the available data. Second, analyses then focused on tenants from Wave 1 who completed both subsequent Waves of the study (N= 814). Bivariate analyses were conducted to compare those who did and did not experience an eviction after Wave 1 on baseline sociodemographic characteristics, COVID-19 status, physical and mental health using chi-square and independent t-tests. Bivariate analyses were then followed with logistic regressions analyses including only significant variables in the bivariate results to identify variables independently predictive of eviction during the eviction moratorium. Third, a preliminary correlation analysis was conducted to examine Wave 1 and Wave 2 variables associated with negative reported effects of the eviction moratorium on tenant-landlord relationships in Wave 3. The correlation analysis was followed by multiple regression analyses including only significant variables found in the correlation analysis to identify independent predictors of negative reported effects of the moratorium on tenant-landlord relationships.

## Results

Table 1 describes the proportions of tenants who were at-risk of eviction, were aware of the eviction moratorium, had delayed paying rent because of the eviction moratorium, and who recently experienced any eviction at each Wave of the study. About 19% of tenants reported they were at-risk for eviction at Wave 1 and this proportion decreased considerable over time over the two subsequent waves. About 7% of tenants at Wave 2 and 3% of Wave 3 tenants reported they had experienced an eviction since follow-up. The majority of tenants were aware of the federal eviction moratorium across the three Waves. Nearly a quarter of Wave 1 tenants reported they delayed paying rent because of the eviction moratorium, which decreased to nearly one-tenth of tenants in Wave 2 to about one-twentieth of tenants at Wave 3. When tenants were asked to rate the effect of the eviction moratorium on the relationship with their landlord, the overall response across waves was neutral to negative in Waves 1, 2, and 3, with a mean rating of 0.03 (SD = 0.77) in Wave 1, rating of -0.01 (SD = 0.50) in Wave 2, and rating of -0.03 (SD = 0.42) in Wave 3 (on a scale from -2 to 2 with negative values indicating a worsened relationship). However, when comparing tenants who did and did not delay paying their rent, those who delayed paying rent were significantly more likely to report a negative effect of the eviction moratorium on the relationship with their landlord.

**Table 1.** Eviction risk and impact during the COVID-19 pandemic (N= 3,393 in Wave 1; N = 1,311 in Wave 2, N = 801 in Wave 3)

| At risk of eviction |  |  |  |
| --- | --- | --- | --- |
|  | Yes, at risk | No, not at risk | N/A |
|  | Raw count<br>(Weighted %)<sup>a</sup> | Raw count<br>(Weighted %) | Raw count<br>(Weighted %) |
| Renters at Wave 1<br>(May-June 2020) | 655 (19.3) | 2605 (76.8) | 133 (3.9) |
| Renters at Wave 2<br>(Sept-Oct 2020) | 97 (7.4) | 1156 (88.2) | 57 (4.3) |
| Renters at Wave 3<br>(Feb-April 2021) | 33 (4.1) | 733 (91.5) | 35 (4.4) |
| Aware there is a moratorium on evictions |  |  |  |
|  | Aware | Not Aware | N/A |
|  | Raw count<br>(Weighted %) | Raw count<br>(Weighted %) | Raw count<br>(Weighted %) |
| Renters at Wave 1 | 2275 (67.0) | 1118 (33.0) | - |
| Renters at Wave 2 | 938 (71.5) | 372 (28.4) | - |
| Renters at Wave 3 | 496 (61.9) | 188 (23.5) | - |
| Delayed paying rent because of eviction moratorium |  |  |  |
|  | Yes | No | N/A |
|  | Raw count<br>(Weighted %) | Raw count<br>(Weighted %) | Raw count<br>(Weighted %) |
| Renters at Wave 1 | 793 (23.4) | 2390 (70.4) | 210 (6.2) |
| Renters at Wave 2 | 123 (9.4) | 1085 (82.8) | 102 (7.8) |
| Renters at Wave 3 | 45 (5.6) | 697 (87.0) | 59 (7.4) |
| Rated effect of eviction moratorium on landlord relationship<sup>b</sup> |  |  |  |
|  | Participants who<br>delayed paying rent<sup>c</sup> | Participants who paid<br>rent on-time | Test of difference |
|  | Mean (SD) | Mean (SD) |  |
| Renters at Wave 1 | 0.02 (1.25) | 0.03 (0.48) | $t(3544) = -0.29^{***}$ |
| Renters at Wave 2 | -0.10 (1.21) | 0.00 (0.38) | $t(1419) = -2.24^{***}$ |
| Renters at Wave 3 | -0.63 (1.11) | 0.01 (0.34) | $t(957) = -10.98^{***}$ |

<sup>a</sup> All percentages shown were calculated as valid percentages.
<sup>b</sup> Effect of eviction moratorium on landlord relationship was rated from -2 (Worsened the relationship greatly) to 2 (Improved the relationship greatly).
<sup>c</sup> Sample sizes for participants who delayed paying rent for each of the Waves was n= 778 for Wave 1, n= 116 for Wave 2, and n= 40 for Wave 3; which was compared to participants who paid their rent on time at each of the Waves which included n= 1,979 at Wave 1, n= 884 at Wave 2, and n= 567 at Wave 3.

| Recently experienced an eviction |  |  |  |
| --- | --- | --- | --- |
|  | Yes | No (or n/a) | N/A |
|  | Raw count<br>(Weighted %) | Raw count<br>(Weighted %) | Raw count<br>(Weighted %) |
| Renters at Wave 2 | 85 (6.5) | 1226 (93.5) | - |
| Renters at Wave 3 | 23 (2.9) | 778 (97.1) | - |

**Note.** \* $p < .05$ , \*\* $p < .01$ , \*\*\* $p < .001$

Among the 814 tenants who completed all three Waves of the study, 35 (4.3%) reported experiencing an eviction after Wave 1. Table 2 shows a comparison between participants who did and did not experience an eviction after Wave 1 on baseline characteristics. Bivariate analyses revealed that participants who experienced an eviction were significantly younger, lower-income, reported a greater number of close friends, and were more likely to be single, black or other race/ethnicity, have dependent children, working full-time, have a history of homelessness, and have reported a negative effect of the eviction moratorium on their tenant-landlord relationship than those who did not experience an eviction.

**Table 2.** Bivariate comparison of Wave 1 demographic, clinical, and psychosocial characteristics of tenants who did and did not experience any eviction after Wave 1 (N = 814)

|  | **Experienced**<br>**eviction at any**<br>**time after Wave 1**<br>**(N = 35)**<br>**Weighted**<br>**Mean/unweighted**<br>**SD or Raw**<br>**n/Weighted%** | **Did not experience**<br>**eviction any time**<br>**after Wave 1**<br>**(N = 779)**<br>**Weighted**<br>**Mean/unweighted**<br>**SD or Raw**<br>**n/Weighted%** | **Test of**<br>**difference**<br>**$t, \chi$** |
| --- | --- | --- | --- |
| **Sociodemographic variables** |  |  |  |
| Age | 36.61 (8.59) | 52.60 (13.42) | $t(1383) = 5.64^{***}$ |
| Gender | | | $\chi^2(1) = 0.64$ |
| Male | 21 (51.3) | 289 (44.8) |  |
| Female | 14 (48.7) | 490 (55.2) |  |
| Race/Ethnicity | | | $\chi^2(6) = 14.52^*$ |
| White non-Hispanic | 20 (52.6) | 536 (70.0) |  |
| Black non-Hispanic | 9 (18.4) | 84 (8.7) |  |
| White Hispanic | 1 (2.6) | 55 (7.7) |  |
| Black Hispanic | 1 (2.6) | 9 (1.3) |  |
| Asian/Pacific Islander | 2 (2.6) | 73 (4.0) |  |
| NA/AN<sup>d</sup> | 0 (0.0) | 1 (0.0) |  |
| Other | 2 (21.1) | 21 (8.3) |  |
| Education | | | $\chi^2(3) = 9.44^*$ |
| High school and below | 5 (12.8) | 75 (8.2) |  |
| Some college | 7 (15.4) | 159 (31.1) |  |
| Associate/Bachelors | 13 (41.0) | 398 (45.0) |  |
| Advanced degree | 10 (30.8) | 147 (15.8) |  |
| Student status | | | $\chi^2(2) = 20.17^{***}$ |
| Not a student | 21 (69.2) | 655 (90.6) |  |
| Part-time | 4 (10.3) | 49 (3.9) |  |

<sup>d</sup> NA/AN= Native American/Alaskan Native

|  |  |  |  |
| --- | --- | --- | --- |
| Full-time | 10 (20.5) | 75 (5.5) |  |
| Marital status | | | $\chi^2(2) = 16.27^{***}$ |
| Single | 15 (55.0) | 315 (30.8) |  |
| D/S/W<sup>e</sup> | 3 (7.5) | 126 (35.7) |  |
| Married/LWP<sup>f</sup> | 17 (37.5) | 338 (33.5) |  |
| # of minors in household | | | $\chi^2(4) = 64.29^{***}$ |
| None | 16 (45.0) | 555 (80.7) |  |
| One | 14 (27.5) | 128 (11.3) |  |
| Two | 3 (12.5) | 67 (5.5) |  |
| Three | 1 (2.5) | 18 (1.6) |  |
| Four or more | 1 (12.5) | 11 (0.8) |  |
| Work Status | | | $\chi^2(4) = 11.32^*$ |
| Full-time | 21 (59.0) | 405 (37.1) |  |
| Half-time | 5 (10.3) | 99 (9.4) |  |
| Unemployed | 5 (12.8) | 107 (11.9) |  |
| R/D/O<sup>g</sup> | 3 (15.4) | 89 (22.2) |  |
| Self-employed | 1 (2.6) | 79 (19.4) |  |
| Personal income | 26178.40<br>(23715.79) | 33850.60<br>(20796.33) | $t(1383) = 2.28^*$ |
| Geographic region | | | $\chi^2(3) = 4.64$ |
| Northeast | 8 (33.3) | 160 (23.9) |  |
| Midwest | 6 (12.8) | 140 (19.3) |  |
| South | 12 (25.6) | 297 (36.5) |  |
| West | 9 (28.2) | 182 (20.3) |  |
| Military veteran | 6 (12.5) | 37 (17.0) | $\chi^2(1) = 0.56$ |
| # of close friends | 1.60 (0.68) | 1.27 (0.27) | $t(1383) = -7.86^{***h}$ |
| Medical Outcomes Study<br>Social Support Survey | 18.06 (5.87) | 19.97 (6.50) | $t(1383) = 1.77$ |
| Any history of homelessness | 20 (57.5) | 79 (10.0) | $\chi^2(1) = 87.42^{***}$ |
| Rating of effect of eviction<br>moratorium on tenant-landlord relationship | -0.17 (1.04) | -0.01 (0.34) | $t(1069) = 2.93^{**}$ |
| **COVID-19 variables** |  |  |  |
| Any positive test for<br>COVID-19 | | | $\chi^2(2) = 41.60^{***}$ |
| Negative | 11 (41.0) | 179 (18.4) |  |
| Positive | 4 (10.3) | 6 (1.0) |  |
| Untested/unknown | 20 (48.7) | 594 (80.6) |  |
| Has anyone close tested<br>positive for COVID-19 | | | $\chi^2(1) = 2.32$ |

<sup>e</sup> D/S/W= Divorced/separated/widowed
<sup>f</sup> LWP= Living with partner
<sup>g</sup> R/D/O= Retired, disabled, others
<sup>h</sup> Significance test for # of close friends was conducted after a log transformation to normalize the variable.

|  |  |  |  |
| --- | --- | --- | --- |
| Positive | 10 (23.1) | 136 (14.3) |  |
| Negative | 25 (76.9) | 643 (85.7) |  |
| **Physical and mental health conditions** |  |  |  |
| # of medical conditions | 1.53 (1.26) | 1.98 (1.64) | $t(1383) = 1.58$ |
| History of psychiatric disorders |  |  |  |
| Schizophrenia-spectrum | 3 (5.1) | 4 (0.2) | $\chi^2(1) = 25.33^{***}$ |
| Posttraumatic stress disorder | 7 (12.8) | 75 (8.7) | $\chi^2(1) = 0.80$ |
| Alcohol use disorder | 6 (12.8) | 26 (2.5) | $\chi^2(1) = 15.26^{***}$ |
| Bipolar disorder | 8 (17.9) | 35 (3.6) | $\chi^2(1) = 19.98^{***}$ |
| Anxiety | 11 (30.8) | 246 (23.4) | $\chi^2(1) = 1.14$ |
| Major depressive disorder | 8 (15.4) | 137 (14.1) | $\chi^2(1) = 0.05$ |
| Drug use disorder | 1 (2.5) | 14 (1.4) | $\chi^2(1) = 0.32$ |
| Traumatic brain injury | 1 (2.6) | 7 (0.7) | $\chi^2(1) = 1.90$ |
| Positive screen for COVID-19 era-related stress symptoms | 21 (46.2) | 84 (7.1) | $\chi^2(1) = 77.24^{***}$ |
| Positive screen for major depression | 24 (69.2) | 228 (24.8) | $\chi^2(1) = 38.75^{***}$ |
| Positive screen for generalized anxiety disorder | 22 (61.5) | 227 (23.4) | $\chi^2(1) = 29.82^{***}$ |
| Past 2-week suicidal ideation | 23 (53.8) | 123 (11.0) | $\chi^2(1) = 64.96^{***}$ |
| Positive screen for alcohol use disorder | 17 (46.2) | 229 (32.4) | $\chi^2(1) = 3.25$ |
| Any illicit drug use in past month | 11 (22.5) | 107 (11.4) | $\chi^2(1) = 4.65^*$ |

**Note.** \* $p < .05$ , \*\* $p < .01$ , \*\*\* $p < .001$

Compared to participants who did not experience an eviction after Wave 1, participants who did experience an eviction were more likely to have been tested for COVID-19. Upon closer examination of only participants who had been tested for COVID-19 (i.e., excluding untested/unknown participants), 20.0% of participants who experienced an eviction reported a positive COVID-19 test at Wave 1 which was significantly higher than the 2.2% of positive COVID-19 participants who did not experience an eviction, χ^2^ (1)= 19.65, p<.001.

Participants who experienced an eviction reported significantly fewer medical conditions at Wave 1 but were more likely to report a history of various psychiatric disorders and more likely to screen positive for current mental health and substance abuse problems at Wave 1, including major depression, generalized anxiety disorder, suicidal ideation, alcohol use disorder, and recent illicit drug use.

These bivariate analyses were followed by multivariable analyses that only included significant variables in the bivariate results. As shown in Table 3, the multivariable analyses revealed participants with a history of homelessness were nearly 9 times more likely to have been evicted than those with no history of homelessness; and participants who reported a greater number of close friends were also more likely to have been evicted as well. COVID-19 status and other clinical variables were not found to be significantly associated with eviction. When these analyses were repeated on only participants who had been tested for COVID-19 (i.e., excluding untested/unknown), history of homelessness remained a major predictor (OR= 9.40, 95% CI= 2.71-32.56, p<.001) but number of close friends was no longer a significant predictor. In addition, having a positive screen for major depression (OR= 7.93, 95% CI= 2.01-31.35, p<.01) and a more negative rating of tenant-landlord relationship (OR= 0.30, 95% CI= 0.11-0.79) was significantly associated with being evicted after Wave 1.

**Table 3.** Logistic regression of tenant characteristics associated with eviction after Wave 1 (N = 814)

| Wave 1 characteristics | Any eviction after Wave 1<br>Odds ratio (95% confidence interval) |
| --- | --- |
| Age | 1.00 [0.93, 1.01] |
| Male | 1.67 [0.70, 3.95] |
| White | 0.95 [0.39, 2.30] |
| Student status | 0.74 [0.22, 2.54] |
| Never married | 1.46 [0.53, 4.03] |
| Any minors in household | 1.20 [0.44, 3.28] |
| Work Status: Employed | 0.81 [0.32, 2.06] |
| # of close friends | 3.26 [1.20, 8.86]* |
| Any positive test for COVID-19 | 2.48 [0.47, 13.18] |
| # of medical conditions | 0.76 [0.53, 1.10] |
| Any history of homelessness | 8.97 [3.43, 23.48]*** |
| Rating of tenant-landlord relationship | 0.75 [0.39, 1.43] |
| Any psychiatric history of mental illness<sup>a</sup> | 0.83 [0.31, 2.22] |
| Positive screen for COVID-19 era-related stress symptoms | 1.97 [0.66, 5.87] |
| Positive screen for major depression | 2.71 [0.91, 8.03] |
| Positive screen for generalized anxiety disorder | 0.74 [0.23, 2.36] |
| Past 2-week suicidal ideation | 2.17 [0.74, 6.37] |
| Positive screen for alcohol use disorder | 0.63 [0.24, 1.64] |

**Note:** Reference groups were tenants who were not white, not students, not employed, married, and tested negative for COVID-19. A log transformation was conducted on number of close friends to normalize the distribution. For the overall model, Nagelkerke $R^2=.40$ , $\chi^2(18)=106.36$ , $p<.001$ . \* $p<.05$ , \*\* $p<.01$ , \*\*\* $p<.001$
<sup>a</sup> Any psychiatric history of mental illness included history of schizophrenia-spectrum disorder, posttraumatic stress disorder, bipolar disorder, anxiety disorder, major depressive disorder, and substance use disorder.

A preliminary correlation analysis found a number of sociodemographic, clinical, and psychosocial variables significantly associated with negative reported effects of the eviction moratorium on tenant-landlord relationships and these variables were entered into a multiple regression analysis (Table 4). The multiple regression analysis found that being female, having a history of psychiatric disorder, recent illicit drug use, and delaying paying rent because of the eviction moratorium were all independently and significantly predictive of a negative reported effect of the eviction moratorium on tenant-landlord relationships. The total R^2^ was .XXX.

**Table 4.** Multiple regression of Wave 1 and 2 tenant characteristics associated with rating of effect of eviction moratorium on tenant-landlord relationships at Wave 3 (N = XXX)^b^

|  | **Wave 3 rating of effect of eviction moratorium on tenant-landlord relationship** |  |  |
| --- | --- | --- | --- |
|  | **Standardized beta coefficient** | **Unstandardized beta coefficient (95% confidence interval)** | **p-value** |
| **Wave 1 variables** |  |  |  |
| Age | .055 | .001 (.000, .003) | .128 |
| Male | .070 | .055 (.002, .107) | .040 |
| Education level | .067 | .016 (.000, .062) | .052 |
| Personal income | .036 | .000 (.000, .000) | .304 |
| Any psychiatric history of mental illness<sup>c</sup> | -.101 | .029 (-.139, -.024) | .005 |
| **Wave 2 variables** |  |  |  |
| Positive screen for COVID-19 era-related stress symptoms | -.055 | .047 (-.167, .018) | .116 |
| Any illicit drug use in past month | -.152 | .042 (-.271, -.105) | <.000 |
| Delayed paying rent because of eviction moratorium | .115 | .045 (.064-.239) | .001 |

<sup>b</sup> Only independent variables significantly correlated with Wave 3 tenant-landlord relationship in preliminary bivariate correlational analyses were included in the multiple regression model.
<sup>c</sup> Any psychiatric history of mental illness included history of schizophrenia-spectrum disorder, posttraumatic stress disorder, bipolar disorder, anxiety disorder, major depressive disorder, and substance use disorder.

## Discussion

In our national sample, we found that the majority (62-72%) of middle- and low-income tenants were aware of the eviction moratorium during the COVID-19 pandemic (May 2020-April 2021). Only a minority (4-19%) of participants reported they were at-risk for eviction across three waves of data collection and 4.3% reported they were evicted over this time period. These findings point to the broad influence of eviction moratorium but also to the reality that some individuals were still evicted despite the moratorium. This may be due to the fact that tenants needed to meet certain qualifications and submit a completed CDC declaration form to their landlord to be protected by the moratorium (Centers for Disease Control and Prevention, 2021a). In addition, the eviction moratorium only prevented evictions due to non-payment of rent and tenants could still be evicted for other legal reasons.

Importantly, substantial proportions of tenants reported they delayed paying their rent because of the eviction moratorium, presumably because they were less concerned about being evicted because of non-payment of rent. However, the federal eviction moratorium expired July 31, 2021 and any unpaid rent or late fees have accumulated as debt that is due at the expiration of the eviction moratorium. Tenants who are unable to pay their accumulated debt may be eligible to be evicted after the moratorium. Notably though, the CDC issued a new targeted eviction moratorium on August 3, 2021 that will remain in effect until October 3, 2021 for “communities with substantial or high levels of community transmission of COVID-19” as designated at the county-level by the CDC (Centers for Disease Control and Prevention, 2021d). Counties with high COVID-19 transmission levels that reduce them below threshold for at least 14 days may have their eviction moratorium lifted before October. Some local jurisdictions have extended the eviction moratorium in their region (San Francisco Rent Board, 2021) so there will be regional variability across the country where the eviction moratorium is in effect and where it is not. Regardless, there is financial assistance available for tenants at-risk for eviction with $21.6 billion dollars in Emergency Rental Assistance funds available through the American Rescue Plan Act (U.S. Department of the Treasury, 2021).

Tenants who reported delayed paying their rent were more likely to report the eviction moratorium had a negative effect on the relationship with their landlords than tenants who did not delay paying rent. There were several other factors associated with the moratorium having a negative effect on tenant-landlord relationships including being female, having a history of psychiatric disorder, and recent illicit drug use. The moratorium may have increased tensions between tenants and landlords particularly among those with mental health and substance abuse problems. This finding would be consistent with research that has shown the COVID-19 pandemic has exacerbated pre-existing mental health problems for some individuals (Muruganandam et al., 2020; Tsai et al., 2021).

While sociodemographic characteristics, history of psychiatric disorder, and current mental health problems were found to be associated with experiencing eviction in bivariate analyses, these associations were no longer significant in multivariable analyses. The strongest predictor of eviction during the eviction moratorium was a history of homelessness, which was not surprising given past housing and financial instability is a known risk factor for eviction (Tsai & Huang, 2019). Additionally, current depressive symptoms, a negative reported effect of the moratorium on your tenant-landlord relationship, and a greater number of friends were also predictive of eviction during the study. Tenants who reported being evicted were much more likely to report a positive COVID-19 test at a 10:1 ratio which supports simulated statistical models that have found evictions can lead to major increases in COVID-19 infection (Nande et al., 2021). It is worth noting though that we did not find COVID-19 was a significant independent predictor of eviction net of other factors so there are many other mediating factors that explain the association between eviction and COVID-19 infection that need further study to elucidate.

## STUDY LIMITATIONS

Likely to have lost people who were evicted; if you lose your home, you’re probably *not* thinking about taking a survey, so it’s like there is something akin to a “survival” bias.

## Data Availability

Data can be requested and made available from the corresponding author.

